# The MultimorbiditY COllaborative Medication Review And DEcision Making (MyComrade) study: A protocol for a cross-border pilot cluster randomised controlled trial

**DOI:** 10.1101/2021.09.16.21263674

**Authors:** Lisa Hynes, Andrew W Murphy, Nigel Hart, Collette Kirwan, Sarah Mulligan, Claire Leathem, Laura McQuillan, Marina Maxwell, Emma Carr, Scott Walkin, Caroline McCarthy, Colin Bradley, Molly Byrne, Susan M Smith, Carmel Hughes, Maura Corry, Patricia M Kearney, Geraldine McCarthy, Margaret Cupples, Paddy Gillespie, John Newell, Liam Glynn, Alberto Alvarez-Iglesias, Carol Sinnott

## Abstract

**Background:** While international guidelines recommend medication reviews as part of the management of multimorbidity, evidence on how to implement reviews in practice in primary care is lacking. The MyComrade (MultimorbiditY Collaborative Medication Review And Decision Making) intervention is an evidence-based, theoretically-informed novel intervention which aims to support the conduct of medication reviews for patients with multimorbidity in primary care. Our aim in this pilot study is to evaluate the feasibility of a trial of the intervention with unique modifications accounting for contextual variations in two neighbouring health systems (Republic of Ireland (ROI) and Northern Ireland (NI)).

**Methods:** A pilot cluster randomised controlled trial will be conducted, using a mixed methods process evaluation to investigate the feasibility of a trial of the MyComrade intervention. A total of 16 practices will be recruited (eight in ROI; eight in NI) and four practices in each jurisdiction will be randomly allocated to intervention or control. Twenty people living with multimorbidity and prescribed ≥10 repeat medications will be recruited from each practice prior to practice randomisation. In intervention practices, the MyComrade intervention will be delivered by pairs of GPs in ROI, and a GP and Practice Based Pharmacist (PBP) in NI. The GPs/GP and PBP will schedule time to review medications together using a checklist. Usual care will proceed in practices in the control arm. Data will be collected via electronic health records and postal questionnaires at recruitment, and 4- and 8-months after randomisation. Qualitative interviews to assess the feasibility and acceptability of the intervention, and explore experiences related to multimorbidity management will be conducted with a purposive sample of GPs, PBPs, practice administration staff and patients in intervention and control practices. The feasibility of conducting a health economic evaluation as part of a future definitive trial will be assessed.

**Discussion:** The findings of this pilot study will assess the feasibility of a trial of the MyComrade intervention in two different health systems. Evaluation of the progression criteria will guide the decision to progress to a definitive trial and inform trial design. The findings will also contribute to the growing evidence-base related to intervention development and feasibility studies.

**Trial registration:** Registry: ISRCTN, ISRCTN80017020; Date of confirmation 4/11/2019; Retrospectively registered; http://www.isrctn.com/ISRCTN80017020.

## Introduction

Multimorbidity, the co-occurrence of two or more long-term conditions, is frequently encountered by general practitioners (GPs), with approximately one in four adults living with multimorbidity, and half of older adults diagnosed with three or more chronic conditions internationally (1). Prescribing is one of the most complex and important considerations of multimorbidity management. Polypharmacy describes the prescription of multiple medications, but consensus is lacking on the threshold number of medications that should be used to define polypharmacy. Although five or more medications is commonly used, more recent studies have used a cut off of ten or more medications to indicate patients at greater risk of adverse events from their medications (2, 3). Certainly, higher numbers of medications are associated with greater risk of preventable drug-related morbidity (4, 5) and the use of multiple medications may impose a burden on individuals, reducing medication adherence (6). The prevalence of polypharmacy is strongly associated with the number of conditions a person has, for example, a large primary care study conducted in Scotland showed 42% of patients with six or more conditions were prescribed 10 or more medications (7). However, using multiple medications for the control of chronic disease may also benefit the patient by reducing morbidity and improving quality of life. A range of factors have been identified as contributing to the complexity of prescribing for general practitioners (GPs) in the context of multimorbidity and the resultant challenges in clinical decision-making (8). For example, specialists initiate many of the medications taken by patients with multimorbidity, but the responsibility for repeat prescribing of these medications lies with primary care (6). Several studies show the dilemmas experienced by GPs who query the ongoing appropriateness of repeat medications, which is further complicated by suboptimal communication between primary and secondary care (9, 10). In addition, drawing on treatment guidelines for prescribing decisions in multimorbidity is often unhelpful or counterproductive as guidelines are designed for single diseases (8).

The prevalence of multimorbidity and polypharmacy continues to grow (11). Yet, there is a lack of intervention research in this area to guide effective management of medications in primary care. (12). Individual structured medication reviews are recommended by the National Institute for Health and Care Excellence (NICE) as key in the management of multimorbidity (13), but evidence shows such reviews frequently do not occur (14). Medication review is a modifiable and potentially impactful behavioural target in multimorbidity management in primary care. The MyComrade (MultimorbiditY Collaborative Medication Review And Decision Making) intervention was developed to address barriers to medication reviewing by GPs, through a series of studies conducted according to the Medical Research Council (MRC) Framework for the development and evaluation of complex interventions, and using the Behaviour Change Wheel (15-17). The COM-B (Capability, Opportunity, Motivation – Behaviour) model and Behaviour Change Technique (BCT) Taxonomy (18) are key features of the Behaviour Change Wheel system, utilised in the development of this intervention. The development and key features of MyComrade are outlined in Table 1 and described in detail elsewhere (15).

**Table 1.** MyComrade intervention: Barriers to medication reviewing and operationalisation of MyComrade BCTs

| Behavioural analysis using COM-B: What are the barriers to medication review in multimorbidity? |  | Selected Behaviour Change Technique and definition | Original operationalisation of BCTs in the MyComrade intervention | Revised operationalisation of BCTs in the MyComrade intervention, based on findings of feasibility study (19) and contextual requirements |
| --- | --- | --- | --- | --- |
| Uncertainty about most valuable medications | Psychological Capability | 3.2 Social support – practical<br>*Advise on, or provide practical help (e.g. colleagues) for performance of behaviour.* | Two GPs collaboratively conduct medication review | Two GPs, or a GP and PBP collaboratively conduct medication review |
| Perceptions related to patient unwillingness to stop long-term medications | Social Opportunity |  |  |  |
| Lack of time to properly review medications | Physical Opportunity | 12.2 Restructuring social environment<br>*Change, or advise to change the social environment in order to facilitate performance of the wanted behaviour.*<br>1.4 Action planning<br>*Prompt detailed planning of performance of behaviour (must include at least one of frequency, context, duration, intensity).* | Plan active medication reviews | Plan active medication reviews |
| An instinct to maintain status quo | Automatic Motivation | 7.1 Prompts/cues<br>*Introduce environmental or social stimulus for the purpose of prompting or cueing behaviour.* | Use of prescribing checklist | Use of prescribing checklist |
| Fear of negative consequences | Reflective motivation | 10.7 Self-incentive<br>*Plan to reward self in future if and only if there has been* | Annual professional appraisal contribution | Annual professional appraisal contribution |
| Opportunity cost of reviewing medications |  |  |  |  |
|  |  | *effort and/or progress in performing the behaviours.* |  |  |
| Based on the findings of the non-randomised feasibility study (19), an additional BCT was added to the MyComrade intervention to provide necessary support to address the opportunity cost of reviewing medications | Reflective motivation | 10.1 Material incentive (Behaviour)<br>*Inform that money, vouchers or other valued objects will be delivered if and only if there has been effort and/or progress in performing the behaviour* |  | Financial reimbursement for review completion<br>*All practices:* €500/£430 for administration associated with recruiting patients.<br>*Intervention practices only:*<br>• €100/£86 for completing the intervention introduction session<br>• €50 /£43 per completed medication review |

The aim of the MyComrade intervention is to support GPs to conduct medication reviews for patients with multimorbidity with a view to optimising the medication regimen and minimising potentially inappropriate prescribing (15-17). As recommended by the MRC framework, the feasibility of MyComrade was tested in a non-randomised qualitative feasibility study (19). The findings showed MyComrade’s acceptability to GPs and that all the medication reviews conducted produced recommendations for medication optimisation. However, participating GPs felt that the sustainability of this approach would require an incentive mechanism, due to the time, personnel demand and opportunity cost of this activity.

The findings of the feasibility study justified proceeding to a randomised pilot trial to address remaining uncertainties and guide the decision to progress to a definitive trial of the intervention (16). In particular, a pilot would allow testing of the MyComrade intervention and study procedures on a small scale (20), and inform important refinements to facilitate the conduct of a robust and transparent definitive trial (16, 21).

A funding opportunity arose for a pilot trial to be conducted across the Republic of Ireland-Northern Ireland border. This opportunity required that unique modifications would be made to the MyComrade intervention to account for contextual differences in the Irish and Northern Irish health systems. The original MyComrade intervention, developed in the Republic of Ireland (ROI), included collaborative medication review by two GPs. In Northern Ireland (NI), the majority of primary care practices have access to a pharmacist, as a result of a Primary Care Pharmacy scheme launched in 2016 (22) (M. Corry, personal communication, June 4, 2020), with a key responsibility for conducting medication reviews (23). Therefore, the MyComrade collaborative medication reviews will take place in Northern Ireland with a GP and a Practice based Pharmacist (PBP). Additionally, based on the findings of the feasibility study (16), the MyComrade intervention was further refined by adding a behaviour change technique (i.e. material incentive) to address the high opportunity cost of medication reviewing.

The aim of this study is to evaluate the feasibility of a trial of the modified MyComrade intervention, including cross-border comparison, using a pilot cluster randomised controlled trial (cRCT). The primary objective is to determine the feasibility of a definitive trial of the MyComrade intervention, focusing on recruitment, retention and fidelity of intervention implementation. The secondary objective is to select suitable outcome and cost effectiveness measures for use in a definitive trial. This study will also enable assessment of the feasibility of MyComrade in two different health systems, producing data on the adaptability and potential generalisability of the intervention.

## Methods

A parallel group, pilot cRCT of the MyComrade intervention will be conducted, where GP practices are the units of randomisation (the clusters), and individual patients with multimorbidity, prescribed 10 or more medications, are the units of analysis (the participants). Figure 1 illustrates the study design. The trial will be conducted based on best practice guidelines for conducting a process evaluation (24) and reported according to the CONSORT guidelines, adapted for pilot studies (21) and SPIRIT (Standard Protocol Items: Recommendations for Interventional Trials) 2013 Statement (25). A completed SPIRIT checklist is provided in Additional file 1.

**Figure 1.**
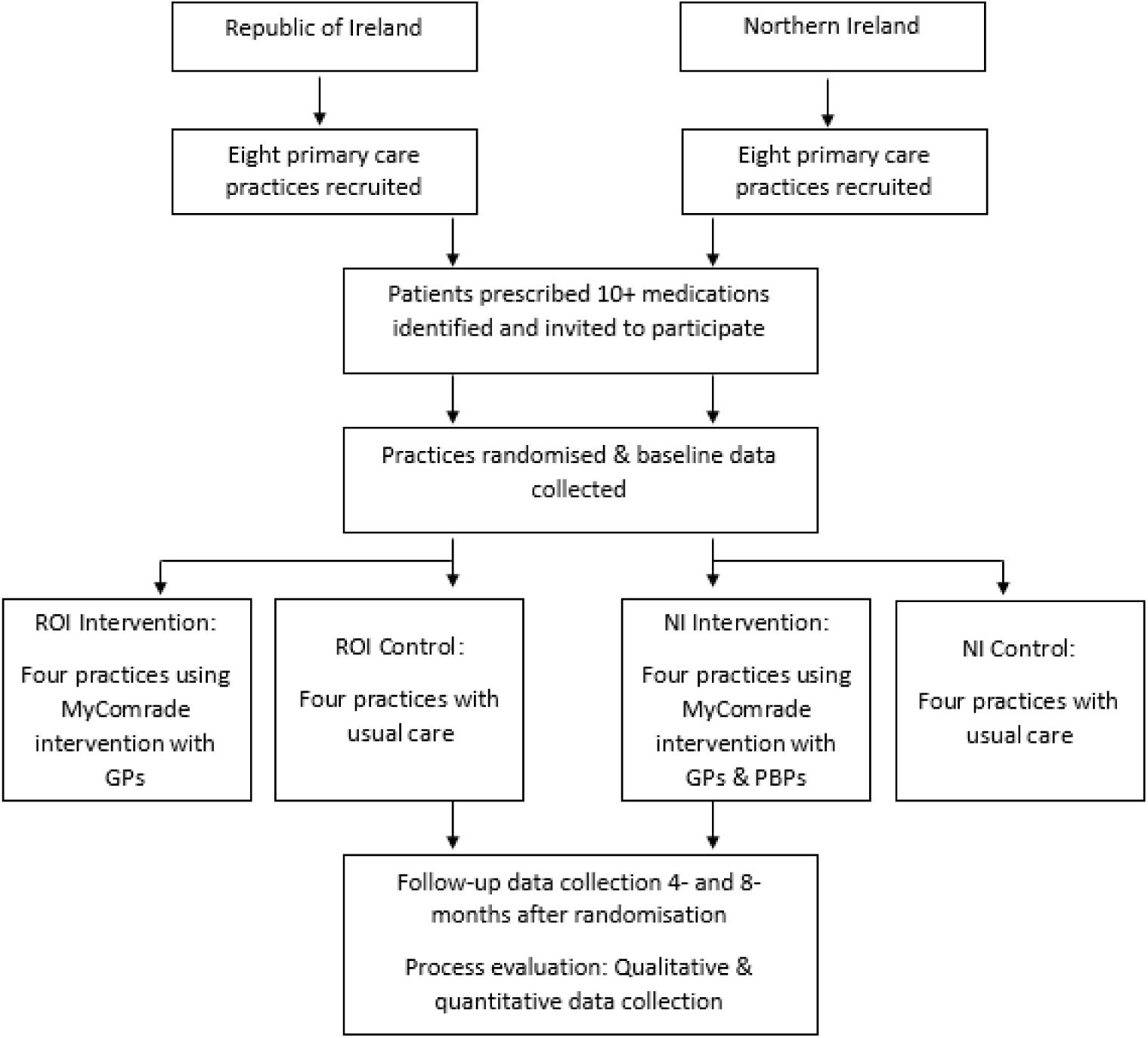
MyComrade study design

A total of 16 practices will be recruited (eight in ROI; eight in NI) and four practices in each jurisdiction will be randomly allocated to the intervention or control arms. Patients with multimorbidity who are prescribed 10 or more repeat prescription medications, will be identified in each practice. From each practice, 20 patients with multimorbidity will be recruited, providing a total of 320 participants. The list of 20 consented patients from each practice will be shared with participating GPs/PBPs. In the intervention arm, pairs of GPs/GP and PBPs will use the MyComrade intervention and will be asked to complete collaborative medication reviews before the 4-month follow-up time point. ROI pairs will be GP-GP and NI pairs will be GP-PBP. Control practices will proceed with usual care. Participant data will be collected from primary care practice records and postal questionnaires sent to participants at baseline before randomisation takes place, and at 4 and 8 months after the date of randomisation. Qualitative interviews to assess the study feasibility and acceptability, and explore experiences related to multimorbidity management will be conducted with a purposive sample of GPs and PBPs participating in the study, practice administration staff, and patients.

A Public and Patient Involvement group has been established, involving four adults (two women and two men) from both NI and ROI and living with multimorbidity. This group have provided input into the patient recruitment materials, and qualitative interview guides, and will provide input into the interpretation of the qualitative findings, and methods for disseminating the study findings. The establishment and running of this group as research partners in this study, is guided by the PPI Ignite @ NUI Galway programme office, part of a national PPI programme funded by the Health Research Board in Ireland.

### Study setting

This study will take place in primary care practices in NI and ROI. The populations of these jurisdictions are similar in terms of ethnicity, with the majority of people in both regions being white (26) and with similar socioeconomic gradients (27). GPs in both jurisdictions work as independent contractors (28) but the health systems differ in important ways, principally that the system in ROI is a mixed public and private system, while the system in NI is publicly funded (29). In ROI, patients are means tested to determine eligibility for a medical card, which entitles them to GP care, medications and other medical services free at the point of access (28). In 2019, 32.4% of the ROI population had a medical card (30). Patients without a medical card pay out of pocket for their medical care and medications. All patients aged over 70 years get free access to GP care but only those with medical cards are eligible for free medications. In NI, GP services are free at the point of access to all patients (31).

Since 2016, most GP practices in NI have access to a PBP, although the hours and role of the PBP will vary depending on the size and specific needs of the practice. Tasks performed by the PBP may include medication reviews and medication reconciliation following discharge from hospital (22).

### Eligibility criteria

#### Primary care practices

Eligible practices in ROI will have at least two GPs willing to conduct medication reviews. In NI, at least one GP and one PBP must be willing to conduct medication reviews for a practice to be eligible. Practices currently involved in other research involving patients with multimorbidity will not be eligible.

#### Patients

Eligible patients must be living with multimorbidity, and prescribed ten or more medications. Patients will not be eligible for this study if they are under 18 years old at the time of medical record review, are pregnant, undergoing terminal illness care, or have cognitive or learning disabilities that would prevent them from completing the study activities.

### Recruitment

Primary care practices will be contacted by the research team via several communication pathways: the NICRN (Northern Ireland Clinical Research Network) Primary Care network; the HRB Primary Care Clinical Trials Network of Ireland (HRB PC CTNI); ROI and NI business directories; local GP education events and meetings; and local GP social media groups. Practices will be sent information on the study and asked to express an interest in taking part. Interested practices will be contacted by a member of the research team to further discuss the study. Before recruitment, each practice will be provided with instructions and asked to run a search in their record system to assess the number of potential participants based on their number of prescription medications (target >60 potentially eligible patients per practice). Practices will be informed at the outset of the material supports associated with participation in the study and details of the implementation of the intervention (Table 1).

Eligible patients will be identified in NI and ROI using electronic record search procedures. These search procedures will be modified to take into account the different electronic practice record systems used in both jurisdictions. In the ROI, a search procedure developed and tested in another Irish primary care multimorbidity trial (32) will be used to identify eligible patients. In NI, a similar search procedure will be developed by the study team and pilot tested for the two main electronic practice record systems there. To comply with General Data Protection Regulation (GDPR), only practice staff members will review the list of eligible patients generated by the search procedure and apply the study eligibility criteria. Eligible patients will receive a recruitment pack, consisting of an invitation to participate in the study, a participant information leaflet, and consent form. Recruitment packs will be provided by the study team, but will be addressed and posted by the practice teams, to adhere to GDPR. To minimise selection bias, patients will be randomly selected from the list of eligible patients and recruitment packs will be sent out until 20 patients from each practice consent to participate.

To limit recruitment bias and help ensure that equal numbers of patients will be recruited in both arms of the trial, randomisation of the practices to intervention or control group will take place after patient recruitment has been completed (33). Practices will be allocated using an online system, called Sealed Envelope, by a biostatistician blinded to allocation. Practices will be allocated by minimisation according to practice size (<4 or 4+ GPs) in the ratio 1:1.

### Intervention

The MyComrade intervention is a complex intervention, which aims to increase the behaviour of active medication review. MyComrade initially involved five components (Table 1); a sixth component material incentive – was added based on results of the first feasibility study of this intervention (19). The intervention is described using the Template for Intervention Description and Replication (TIDieR) Checklist (34) in Additional file 2.

MyComrade will be implemented in intervention practices following recruitment of participants, baseline data collection and practice randomisation. The research team will deliver a brief introduction session on the intervention. The introduction session will be audio-recorded to allow independent fidelity assessment, in terms of content and duration. The GP research fellow with the NI and ROI based study teams (LMQ and SM) will contact each intervention practice 3 and 6 weeks after this session to gauge progress in terms of medication review completion and address any study related questions.

Participating GPs and PBPs will be given the list of eligible patients from their practice who consent to take part. GP/GP and PBP pairs will schedule a time to meet each other to conduct each collaborative medication review, using a medication review checklist to guide the discussion. The medication review checklist was adapted from the “NO TEARS” tool for medication review (35), originally designed for doctor-patient medication reviewing. This seven-item tool was selected due to its simplicity, and was described by GPs in the feasibility study for the MyComrade interventions as a helpful guide for medication review (19). The earlier feasibility study suggested that reviews take between 10 and 30 minutes each. GP/PBPs will scan the completed checklist into the participant’s practice record, highlighting any potential options for medication changes, and discussing these with the participant prior to making changes.

### Progression criteria

Progression criteria for this study (Table 2) were developed based on the Avery et al. Top Ten Tips for guiding the decision to progress from a pilot to a definitive trial, focusing on key acceptability and feasibility variables (36). Several rounds of discussions were held within the study team to draft the criteria outlined in Table 2. Discussions were guided by existing literature, study team experience, the pilot study design, and potential barriers and facilitators to practice and patient involvement. As part of the process evaluation within this study, qualitative and quantitative data will be collected for investigation of the following outcomes: feasibility of practice recruitment, feasibility of patient recruitment, feasibility of practice retention, feasibility of patient retention, and feasibility of intervention implementation.

**Table 2.**
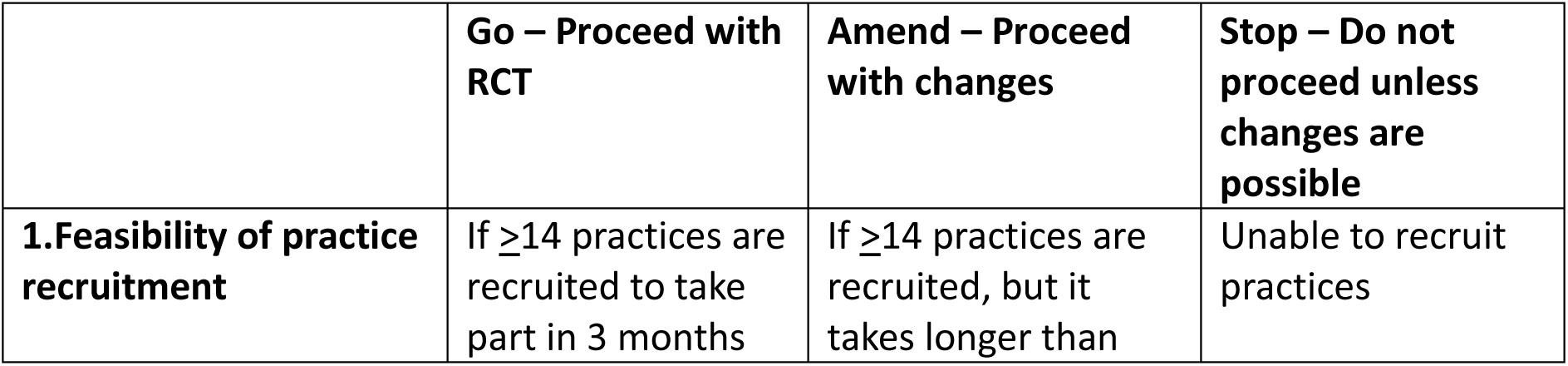
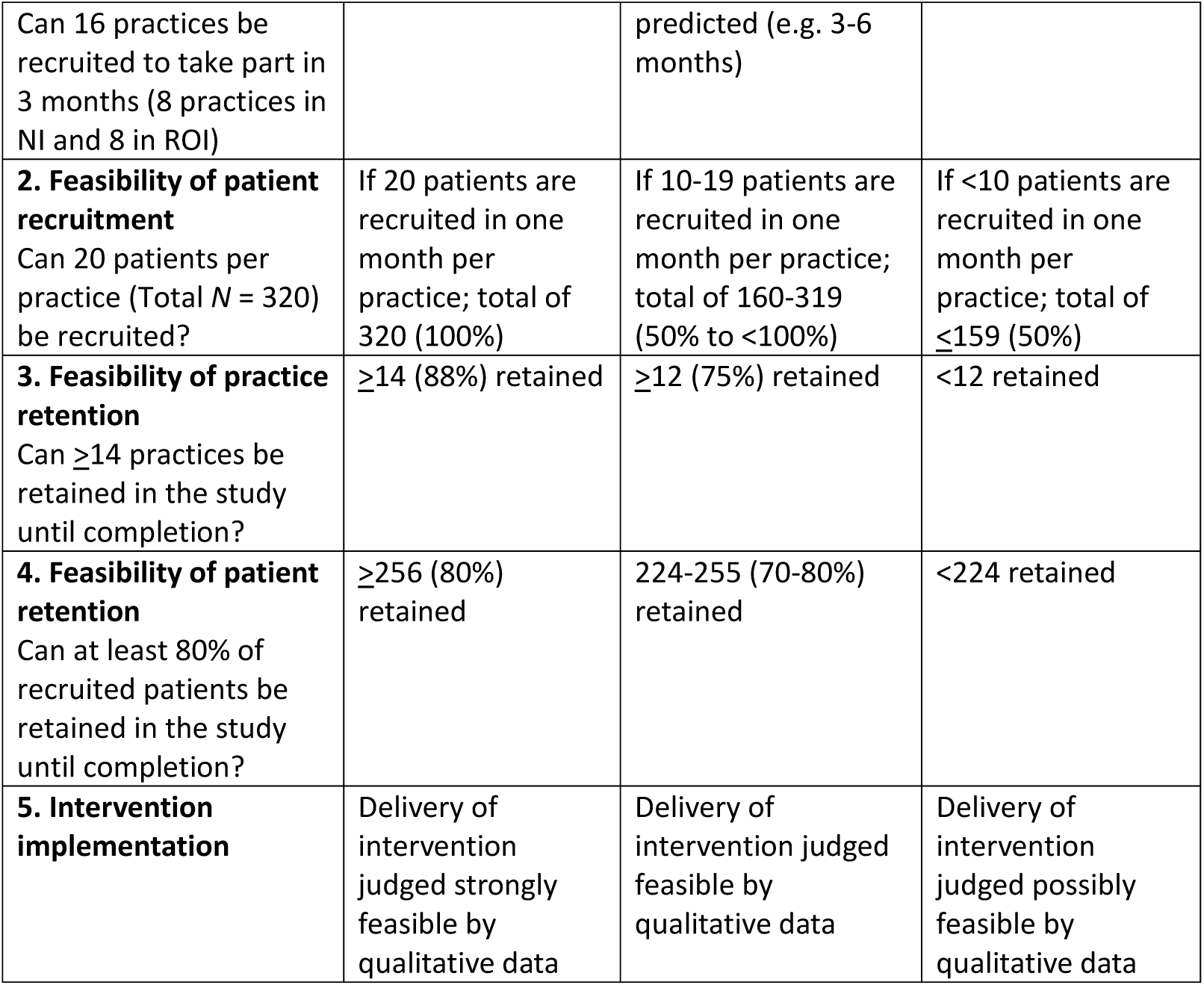
MyComrade Progression Criteria

### Outcomes

The primary outcomes relate to the progression criteria: the feasibility of practice recruitment, patient recruitment, practice retention, patient retention and feasibility of intervention implementation. Secondary outcomes are: completion of medication reviews (GP/PBP outcome); treatment burden and quality of life (patient outcomes); and number of prescribed medications, changes in prescribed medications, deprescribing and indicators of potentially inappropriate prescribing (prescribing outcomes). The indicators of potentially inappropriate prescribing were adapted from the set of evidence-based, validated prescribing safety indicators developed in the PINCER trial (37). The secondary outcomes will be used to inform the choice of primary outcome should we proceed to a definitive trial at a later stage. The logic model in Figure 2 illustrates the intervention components, proposed mechanisms of impact of each component, contextual factors, and outcomes. The logic model was designed based on guidance from Moore et al. (38).

**Figure 2.**
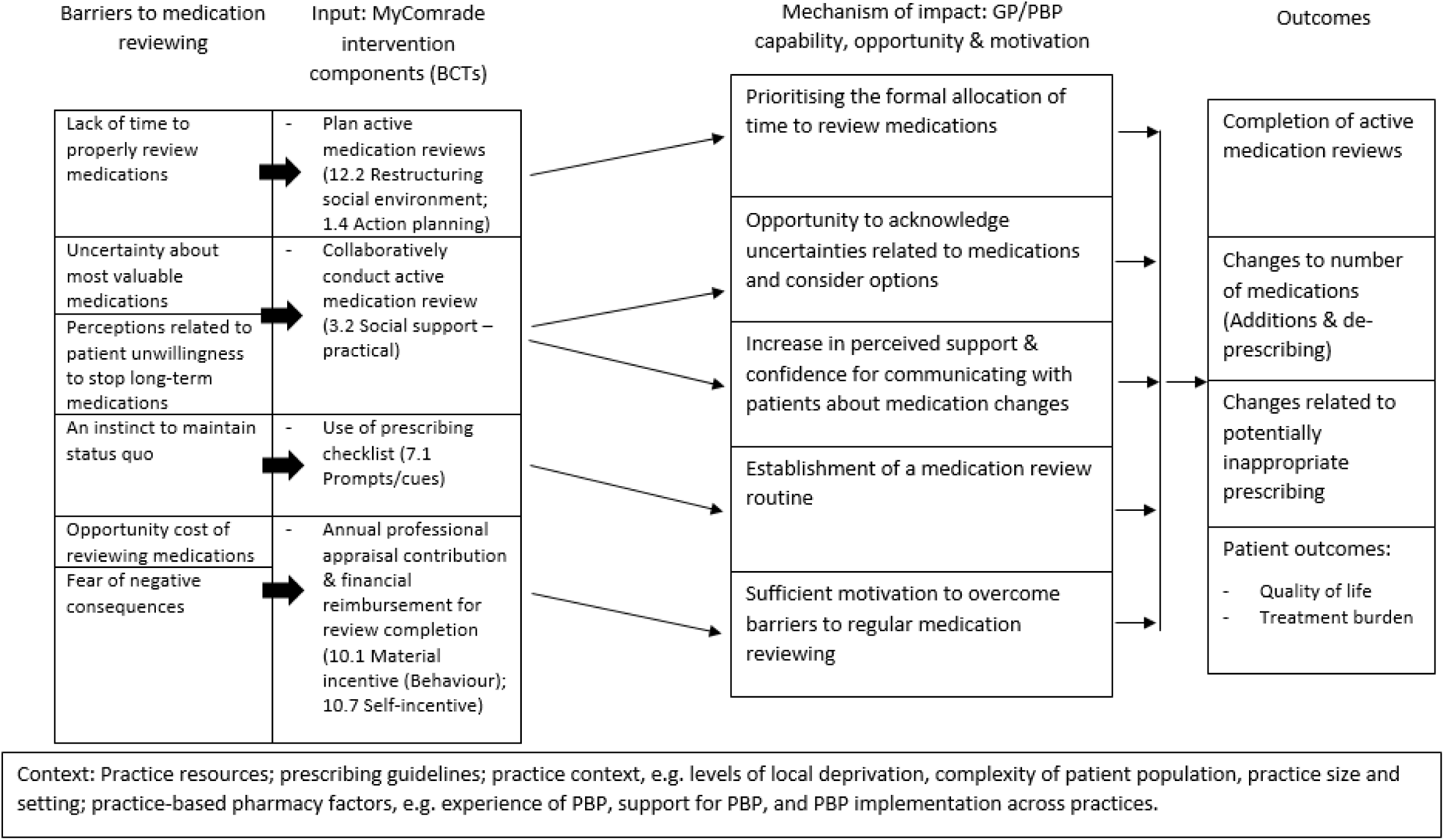
MyComrade logic model

Data will be collected on GP practice demographic information, and patient demographic information and healthcare utilisation. Additional file 3 provides a summary of the variables measured in this study, and the data source for each. The secondary outcomes will be assessed at baseline, and 4- and 8-months after randomisation of practices. The variability, consistency, response rates, success of data collection methods and data completeness for each outcome will be determined to understand the feasibility and acceptability. These findings will help to determine the primary outcome(s) for a future definitive trial.

### Sample size

As this is a pilot study, a formal sample size calculation was not done (39). This study aims to recruit 16 primary care practices (Eight in NI and eight in ROI) and 20 patients per practice (*N* = 320), based on recommendations from Eldridge et al. (20) and the CONSORT guidance (21) on the minimum number of clusters required in pilot and definitive cRCTs respectively. The aims of this pilot cRCT are to investigate feasibility and acceptability, and to identify the most suitable primary outcome(s) for a definitive cluster randomised trial. The estimates from this pilot trial will be used to calculate the sample size needed using the methods outlined by Rutterford et al. (40).

There are approximately 73 primary care practices in the border region where recruitment will take place in ROI, and 330 practices across NI. The average number of publically funded patients registered with a GP in the ROI is 1,124, and over 90% of GPs also see private patients, with the majority reporting 500-1000 private patients (28). In Northern Ireland, practices are more likely to be group practices and may have larger list sizes (41). As we will only recruit practices with at least two full time GPs, we estimate that there will be at least 130 patients potentially eligible for the pilot cRCT in each practice. We will invite 50 patients per practice, and a priori, have defined a success criterion as 40% of the total number of participants invited to be recruited to the research evaluation (approximately 20 patients per practice).

### Process evaluation and qualitative data collection and analysis

The main purpose of the process evaluation is to answer questions relating to the primary feasibility outcomes (recruitment, retention and intervention implementation). The process evaluation is informed by the approach described by Grant et al. (24), and utilises quantitative and qualitative methods across 11 framework domains (described in Table 3). Semi-structured interviews will be used to collect qualitative data from a purposive sample of one GP or PBP, and a practice manager or administrator from each practice. From the patients recruited from each practice, one patient will be invited for an interview, through random sampling. If a patient does not respond or declines to be interviewed, a second patient will be randomly selected and invited for interview. The topic guides will explore the experiences of those participating in the study and issues of implementation following Proctor et al.’s (42) taxonomy of implementation outcomes (acceptability, adoption, appropriateness, feasibility, fidelity, implementation cost, coverage and sustainability– see Table 3). Interviews will be conducted remotely by telephone (by EC and LH) with audio-recording, and last approximately 30-60 minutes. Due to the geographical spread of the study, telephone interviews are most feasible, and this approach has been found to be an acceptable alternative to in-person interviews (43).

**Table 3.**
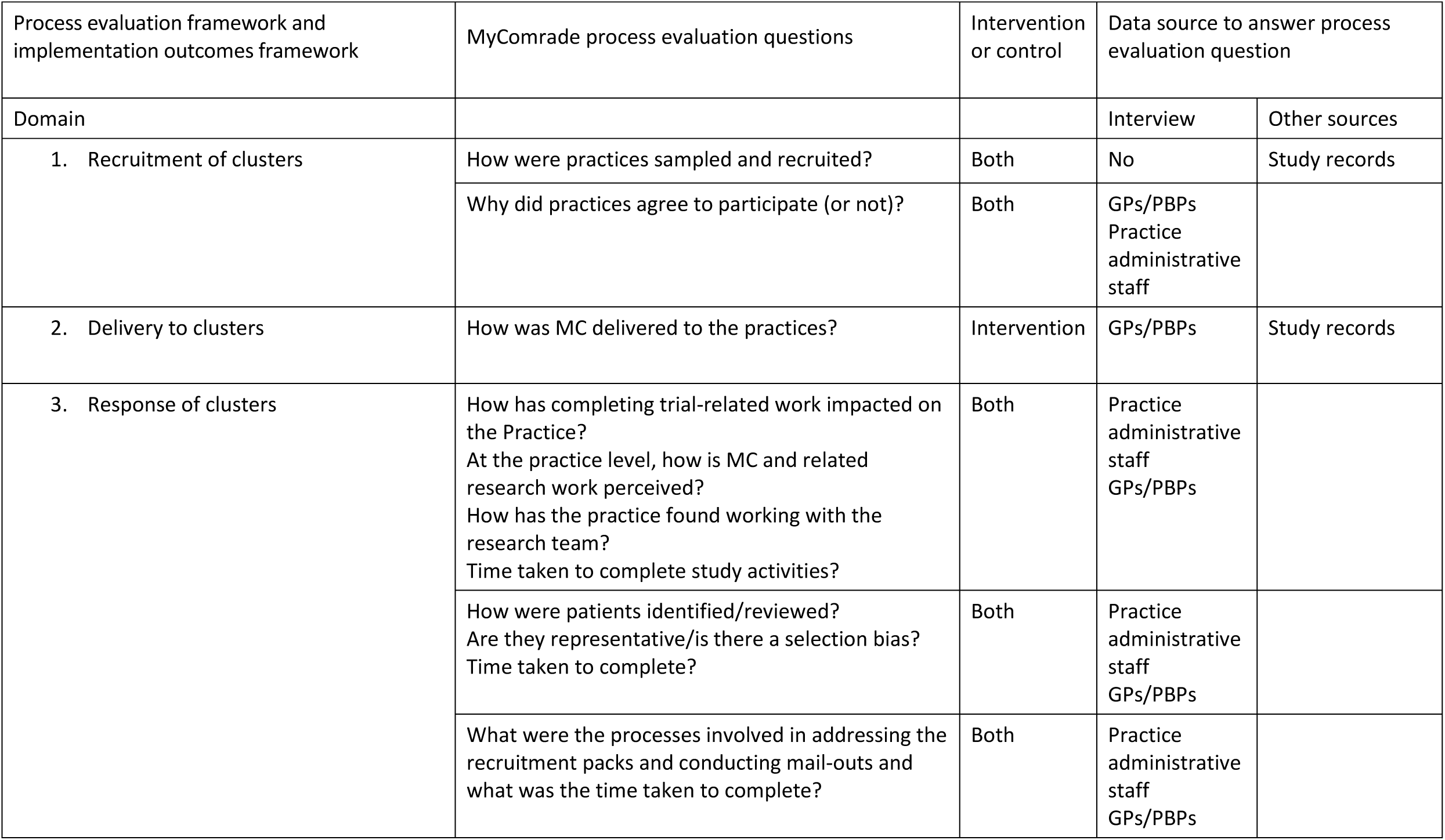
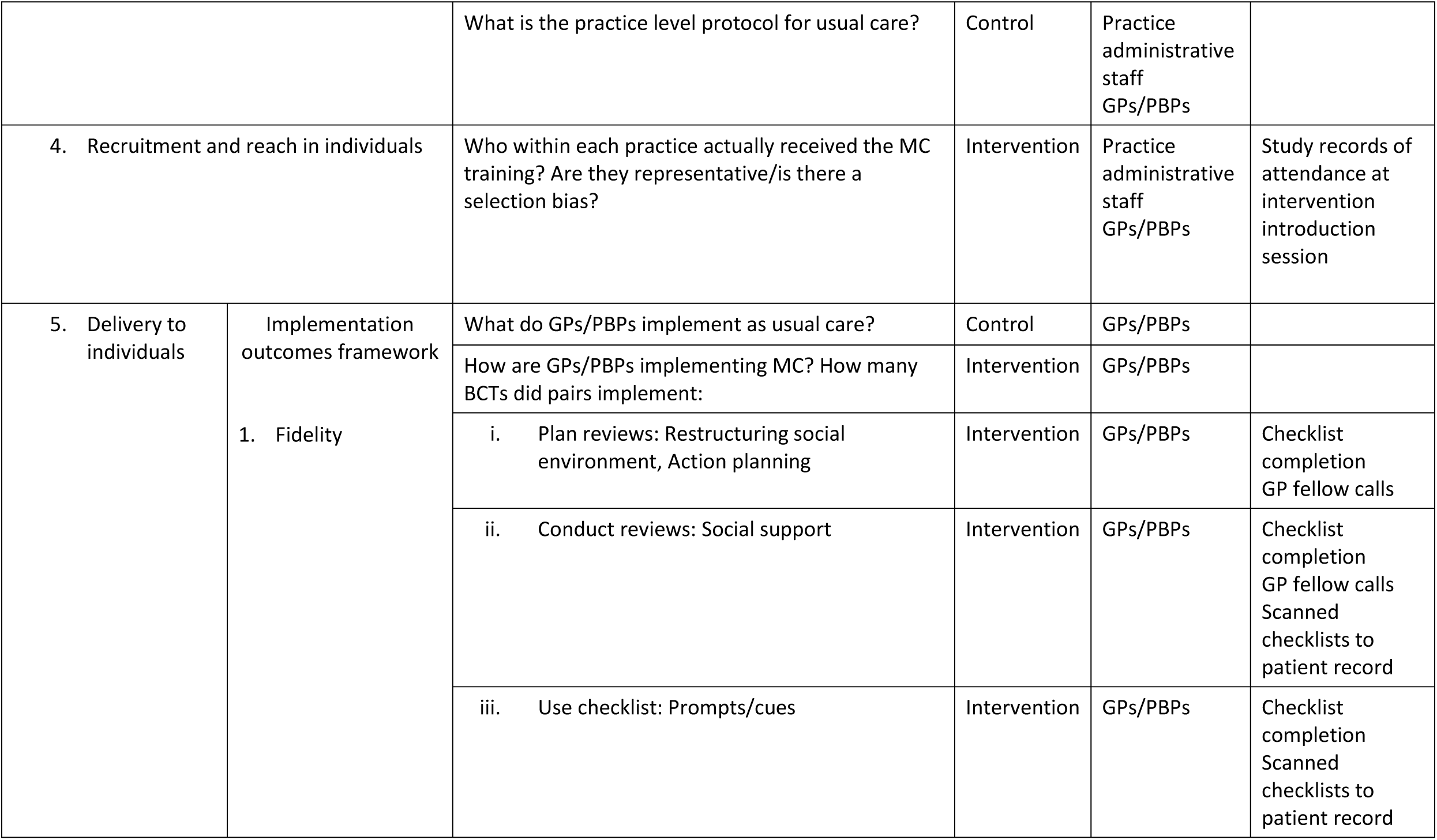
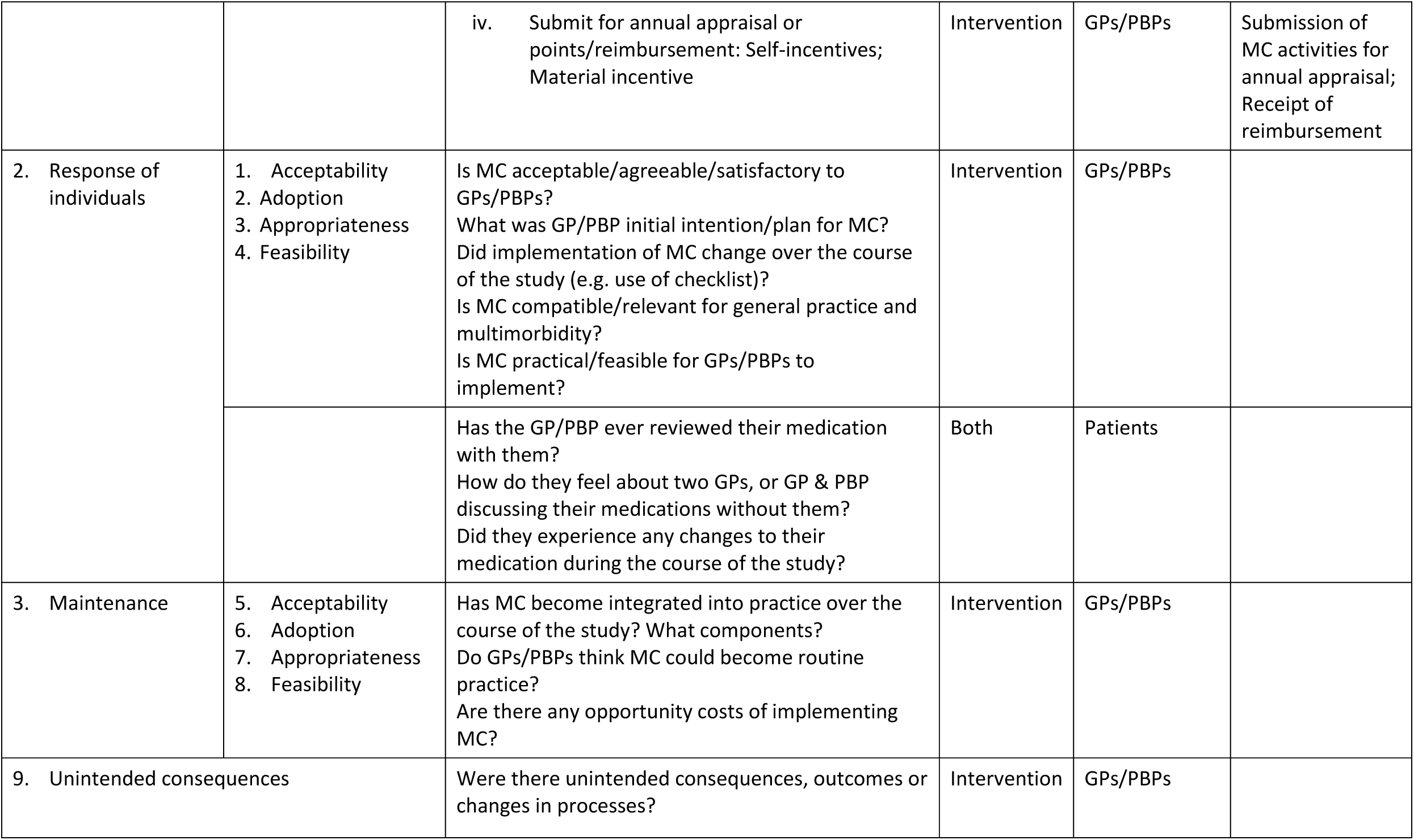
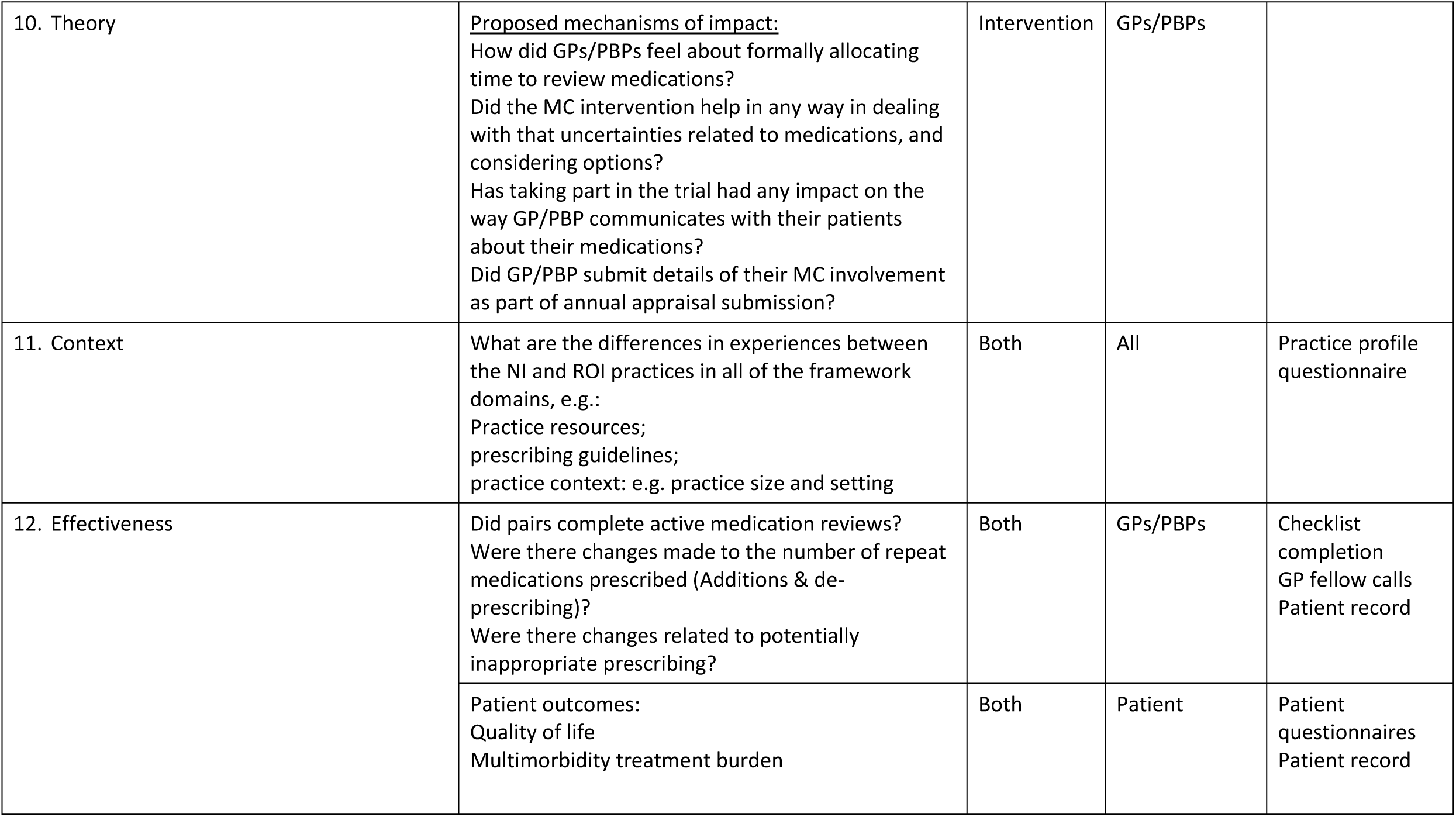
MyComrade study process evaluation framework, including implementation outcomes framework

| Process evaluation framework and implementation outcomes framework | MyComrade process evaluation questions | Intervention or control | Data source to answer process evaluation question |  |
| --- | --- | --- | --- | --- |
| Domain |  |  | Interview | Other sources |
| 1. Recruitment of clusters | How were practices sampled and recruited? | Both | No | Study records |
|  | Why did practices agree to participate (or not)? | Both | GPs/PBPs<br>Practice administrative staff |  |
| 2. Delivery to clusters | How was MC delivered to the practices? | Intervention | GPs/PBPs | Study records |
| 3. Response of clusters | How has completing trial-related work impacted on the Practice?<br>At the practice level, how is MC and related research work perceived?<br>How has the practice found working with the research team?<br>Time taken to complete study activities? | Both | Practice administrative staff<br>GPs/PBPs |  |
|  | How were patients identified/reviewed?<br>Are they representative/is there a selection bias?<br>Time taken to complete? | Both | Practice administrative staff<br>GPs/PBPs |  |
|  | What were the processes involved in addressing the recruitment packs and conducting mail-outs and what was the time taken to complete? | Both | Practice administrative staff<br>GPs/PBPs |  |

|  |  |  |  |  |  |
| --- | --- | --- | --- | --- | --- |
|  |  | What is the practice level protocol for usual care? | Control | Practice administrative staff<br>GPs/PBPs |  |
| 4. Recruitment and reach in individuals |  | Who within each practice actually received the MC training? Are they representative/is there a selection bias? | Intervention | Practice administrative staff<br>GPs/PBPs | Study records of attendance at intervention introduction session |
| 5. Delivery to individuals | Implementation outcomes framework<br>1. Fidelity | What do GPs/PBPs implement as usual care? | Control | GPs/PBPs |  |
|  |  | How are GPs/PBPs implementing MC? How many BCTs did pairs implement: | Intervention | GPs/PBPs |  |
|  |  | i. Plan reviews: Restructuring social environment, Action planning | Intervention | GPs/PBPs | Checklist completion<br>GP fellow calls |
|  |  | ii. Conduct reviews: Social support | Intervention | GPs/PBPs | Checklist completion<br>GP fellow calls<br>Scanned checklists to patient record |
|  |  | iii. Use checklist: Prompts/cues | Intervention | GPs/PBPs | Checklist completion<br>Scanned checklists to patient record |
|  |  | iv. Submit for annual appraisal or points/reimbursement: Self-incentives; Material incentive | Intervention | GPs/PBPs | Submission of MC activities for annual appraisal; Receipt of reimbursement |
| 2. Response of individuals | 1. Acceptability<br>2. Adoption<br>3. Appropriateness<br>4. Feasibility | Is MC acceptable/agreeable/satisfactory to GPs/PBPs?<br>What was GP/PBP initial intention/plan for MC?<br>Did implementation of MC change over the course of the study (e.g. use of checklist)?<br>Is MC compatible/relevant for general practice and multimorbidity?<br>Is MC practical/feasible for GPs/PBPs to implement? | Intervention | GPs/PBPs |  |
|  |  | Has the GP/PBP ever reviewed their medication with them?<br>How do they feel about two GPs, or GP & PBP discussing their medications without them?<br>Did they experience any changes to their medication during the course of the study? | Both | Patients |  |
| 3. Maintenance | 5. Acceptability<br>6. Adoption<br>7. Appropriateness<br>8. Feasibility | Has MC become integrated into practice over the course of the study? What components?<br>Do GPs/PBPs think MC could become routine practice?<br>Are there any opportunity costs of implementing MC? | Intervention | GPs/PBPs |  |
| 9. Unintended consequences |  | Were there unintended consequences, outcomes or changes in processes? | Intervention | GPs/PBPs |  |
| 10. Theory | <u>Proposed mechanisms of impact:</u><br>How did GPs/PBPs feel about formally allocating time to review medications?<br>Did the MC intervention help in any way in dealing with that uncertainties related to medications, and considering options?<br>Has taking part in the trial had any impact on the way GP/PBP communicates with their patients about their medications?<br>Did GP/PBP submit details of their MC involvement as part of annual appraisal submission? | Intervention | GPs/PBPs |  |
| 11. Context | What are the differences in experiences between the NI and ROI practices in all of the framework domains, e.g.:<br>Practice resources;<br>prescribing guidelines;<br>practice context: e.g. practice size and setting | Both | All | Practice profile questionnaire |
| 12. Effectiveness | Did pairs complete active medication reviews?<br>Were there changes made to the number of repeat medications prescribed (Additions & de-prescribing)?<br>Were there changes related to potentially inappropriate prescribing? | Both | GPs/PBPs | Checklist completion<br>GP fellow calls<br>Patient record |
|  | Patient outcomes:<br>Quality of life<br>Multimorbidity treatment burden | Both | Patient | Patient questionnaires<br>Patient record |

Interviews will be transcribed verbatim and analysed using the framework approach (44) in Nvivo software. At the outset, up to six transcripts will be coded inductively by two researchers (LH & CK), who will then meet to discuss initial coding and to agree on an analytical framework. The agreed framework will be refined inductively through subsequent rounds of coding and team discussion. Data will be summarised using a framework matrix of the themes and sub-themes through a process known as charting, which will allow data summaries to be easily generated and linked to relevant data. The two coders and wider study team, including the PPI group, will work together to interpret the findings.

### Quantitative data collection and analysis

Data relating to progression criteria (e.g. recruitment and retention rates) will be collected throughout. Quantitative data will be collected from patients through postal questionnaires, once they have provided informed consent, at baseline, and 4- and 8-months after practice randomisation (Additional file 3). Prescribing data will be collected by research nurses from practice record systems, and will be based on a data collection tool developed and pilot tested in advance (Additional file 3). An intention to treat analysis will be conducted, with data from all eligible patients being included in analyses. We will determine estimates of the variability in secondary outcomes (e.g. treatment burden and health-related quality of life; potentially inappropriate prescribing, number of prescribed medications, and rates of deprescribing; and completion of medication reviews) at baseline and/or study end, the variability in the change in responses over time, and the likely proportion of missingness in the responses. Quantitative data will inform the decisions on the number of clusters required, optimal cluster size and potential intracluster correlation for a subsequent definitive trial.

### Exploratory Analysis

Linear and nonlinear mixed effects models, that account for the cluster design and allow adjustment for baseline measurements, will be used to tentatively explore differences in the secondary outcomes between pilot trial arms.

### Health Economic Analysis

The health economic study will be conducted alongside the pilot cRCT to explore the feasibility of conducting an economic evaluation to assess the cost effectiveness of the MyComrade intervention. Data collection tools will be developed for the purposes of collecting data on resource use and outcome measures over the trial follow-up period. An exploratory process will be conducted to identify the resource use and costs associated with delivery of the intervention, in addition to the costs of clinical actions linked to the medication reviews, and other healthcare resource use by patients. Unit costs will be identified and applied to convert data on resource use to costs. For the pilot cost utility analysis, Quality Adjusted Life-Years (QALYs) will be generated using the EuroQol EQ-5D-5L (45). Preliminary incremental analysis will be undertaken to provide information on the marginal costs and marginal effects of the MyComrade+ Intervention relative to usual practice and a range of techniques will be employed to address uncertainty. Preliminary subgroup analysis will compare data for the two different healthcare settings (46, 47). This analysis is designed to determine feasibility of this approach and not cost effectiveness.

### Data management and protection

A data management plan will be agreed upon by the study teams in ROI and NI. All data will be safeguarded in a manner that meets the requirements of the Data Protection Acts, 1988 and 2003 in ROI, and the Data Protection Act 2018, in NI. Data collected from practice record systems will be anonymised and labelled with participants’ study identification (ID) number before being removed from practices. Questionnaires will also be anonymous, and the only identifier will be the participant’s ID number. To enable communication with participants for follow-up questionnaires and qualitative interviews, study ID numbers and contact information will be stored on a document kept separate from data and signed consent forms.

Qualitative interview data collected from patients, GPs/PBPs and practice staff will be stored securely and labelled using an anonymous study ID number. Audio recordings will be transcribed verbatim and identifying information will be removed.

Data collected from practice record systems will be stored electronically on a secure platform. Participant questionnaire data will be entered into an anonymous study database and stored electronically on a secure platform. Hard copies of source data and data collection forms related to participant practice-level data, as well as participant questionnaires, will be stored securely in a locked cabinet in a locked office for the duration of the study. Audio recordings and transcripts of interviews will be stored electronically on a secure platform, and recordings will be deleted from the recording equipment. Only study team members will have access to the data.

A Trial Steering Committee consisting of an independent chairperson, a GP, pharmacist, health psychologist and two public and patient representatives provides oversight and guidance to the research team regarding protocol implementation and challenges that arise. Major amendments to the protocol will be reported to relevant parties, such as Research Ethics Committees, and will be described in the final published manuscript. Participating practices, patients and PPI group members will receive a summary of the study findings.

### Ethical approval

Ethical approval has been granted by the Irish College of General Practitioners Research Ethics Committee (ROI) and the Office of Research Ethics Committees Northern Ireland (ORECNI).

### Study status

At the time of submission of this study protocol (Version 1.3; date, June 5, 2019) recruitment of primary care practices and patients has been completed. Data collection will be complete in March 2021. Recruitment was completed before the COVID-19 pandemic began and most study activities paused between March and May 2020. Since study activities restarted in June 2020, intervention introduction sessions have been virtual. Data collection has proceeded as originally planned, and research nurses take necessary precautions when entering practices to collect patient data, for example wearing Personal Protective Equipment.

## Discussion

This study will assess the feasibility of a trial of the MyComrade intervention by conducting a pilot cRCT of the intervention in the ROI and in NI. Despite being recommended to reduce the burden and potential for adverse outcomes associated with polypharmacy, medication reviews are often underused in primary care (6). Informal peer support for treatment decision-making is a common and valuable practice in primary care (10). By bringing GPs/GPs and PBPs together, with supportive tools such as an organising checklist and allocated time, MyComrade aims to facilitate the sharing of expertise and experience to overcome the persistent barriers to medication management in primary care. The introduction of PBPs in NI provides a unique opportunity to compare alternative approaches to enhancing medication reviews across different healthcare systems. The testing of this intervention in two different health systems (ROI and NI), will provide data on the adaptability and generalisability of the intervention. Although the populations of NI and ROI are broadly similar, self-reported health has been reported in one study to be lower in NI than in ROI - a feature that will warrant consideration in any future definitive cross-border trial of this intervention (26).

On completion of this study, the MyComrade intervention will have progressed through the Development and Feasibility/piloting phases of the MRC framework (16). The development of interventions using a systematic approach such as that of the MRC framework is widely recommended to address persistent issues with study quality, effectiveness and implementation (48). The theoretical basis for this intervention and specification of proposed mechanisms of impact enable a level of description and testing of the intervention that is now widely called for in intervention research (49). Therefore, the findings of this study with respect to the pre-specified progression criteria and effectiveness outcomes will provide a strong indication of the appropriateness of moving to a full-scale trial.

The purpose of feasibility and piloting, according to the MRC, is to test procedures, estimate recruitment and retention rates and determine sample size (16). The science of feasibility and pilot studies has been developing rapidly in recent years, and an extension to the CONSORT 2010 statement was published in 2016 to address issues in reporting and conduct (21). The feasibility and pilot phase in the development of the MyComrade intervention was designed based on recent definitions produced by Eldridge et al., (20). Using a multi-method approach including a Delphi study and systematic review, Eldridge and colleagues developed a conceptual framework that presents feasibility as the overarching term for studies done in preparation for an RCT. This pilot cRCT was designed as a smaller version of a future full-scale cRCT, with the aim of assessing the feasibility of procedures such as recruitment and randomisation. This study builds on the findings of the previous non-randomised feasibility study of the MyComrade intervention, which focused on the acceptability of the intervention to GPs, its adaptability for GPs working in different contexts, and whether recommendations for optimising medications resulted from the medication reviews.

The overarching aim of this intervention is to optimise the management of medications prescribed for people living with multimorbidity, specifically those with prescriptions for 10 or more repeat medications. This study is an essential step in examining the potential for MyComrade to achieve this overarching aim. To gain an accurate understanding of the complex issues related to polypharmacy and medicines management in multimorbidity, and produce an effective and implementable intervention, this programme of research has been conducted closely in line with current recommendations. Therefore, this research will contribute to the evidence-base related to intervention development and feasibility testing, and the management of multimorbidity in primary care nationally and internationally.

## Data Availability

All data referenced in this protocol are available on request from the corresponding author

## List of abbreviations

cRCT: Cluster Randomised Controlled Trial
GP: General Practitioner
HRB PC CTNI: Health Research Board Primary Care Clinical Trials Network of Ireland
MC: MyComrade
MRC: Medical Research Council
NI: Northern Ireland
NICE: National Institute for Health and Care Excellence
NICRN: Northern Ireland Clinical Research Network
ORECNI: Office of Research Ethics Committees Northern Ireland
PBP: Practice Based Pharmacist
QALYs: Quality Adjusted Life-Years
ROI: Republic of Ireland
TIDieR: Template for Intervention Description and Replication

## Declarations

Ethics approval and consent to participate: Ethical approval has been granted by the Irish College of General Practitioners Research Ethics Committee (ROI) and the Office of Research Ethics Committees Northern Ireland (ORECNI; REC reference 19/NI/0120; IRAS ID 260425). Informed consent was obtained before recruiting participants.

### Consent for publication

NA

### Availability of data and materials

NA

### Competing interests

The authors declare that they have no competing interests.

### Funding

HSC R&D Division Cross-border Healthcare Intervention Trials in Ireland Network (CHITIN), UK and Ireland. The views and opinions expressed in this protocol do not necessarily reflect those of the European Commission or the Special EU Programmes Body (SEUPB).

### Authors’ contributions

LH prepared the manuscript with input from all authors; All authors read and approved the final manuscript; CS developed and pilot tested the intervention with input from CB and MB.

### Trial sponsor

National University of Ireland, Galway;

## Acknowledgements

The authors would like to thank the primary care practices who have joined the MyComrade study, and all patients who have agreed to participate. The authors would also like to acknowledge the invaluable input of the MyComrade Public and Patient Involvement group into the conduct of the study to date.

## Notes

### Competing Interest Statement

The authors have declared no competing interest.

### Clinical Trial

ISRCTN80017020

